# Individual progression of S100 calcium binding protein beta as a surrogate for epilepsy risk (PROG-S100B): rationale and design of a meta-analysis project

**DOI:** 10.1101/2025.10.28.25338942

**Authors:** Yan Bo

## Abstract

**Background:** Epilepsy research centres in Europe and Asia have reported the correlation between S100 Calcium Binding Protein beta (S100B) and epilepsy. However, whether S100B has clinical diagnostic value has been debated. This epilepsy study developed a long-term research plan to resolve this controversy, divided into three simple phases.

**Methods:** The first phase is mainly observational studies, focusing on resolving the debate on the correlation between S100B levels in serum or cerebrospinal fluid and epilepsy. The second phase uses observational and randomized controlled research data, focusing on solving the baseline standard of S100B level and epilepsy population with different characteristics and providing a research basis for the predictive value of S100B level on epilepsy. The third phase is mainly based on randomized controlled studies, focusing on the accuracy of S100B as a biomarker for diagnosing epilepsy.

**Discussion:** Due to the large number of high-risk groups for epilepsy, it is difficult to identify whether patients with involuntary jerks of limbs have epilepsy. Although electroencephalogram (EEG) can capture pathological brain waves, it inconveniences the general population. Even when a patient is diagnosed with epilepsy and discharged from the hospital, predicting the time interval for the subsequent seizure is difficult. Therefore, a more convenient and clinically valuable tool than the EEG is necessary. The number of studies on epilepsy biomarkers has gradually increased in recent years, which provides a sufficient research basis for this study.

**Registration:** https://www.crd.york.ac.uk/PROSPERO/, CRD-ID: CRD42023425431.

## 1 Introduction

The gene encoding S100B is on 21q22.3, which changes gene expression(1) associated with neurological diseases (**Figure 1**) such as epilepsy (www.genecards.org). The expression level of S100B can now be detected by serum or cerebrospinal fluid. In studying brain injury biomarkers, S100B is often considered a significant factor associated with neurological diseases such as epilepsy(2) and melanoma(3)(www.genecards.org). Among these findings, S100B was considered a potential predictive marker for epilepsy. However, these findings did not reach the endpoint. Is the result that the expression level of S100B is associated with epilepsy reliable? What kind of baseline can S100B establish as a diagnostic marker for epilepsy? How does the S100B level serve as an evaluation for treating epilepsy in one day, week, or month? The debate on the relevance of S100B expression levels to epilepsy will end in this long-term research program.

**Figure 1.**
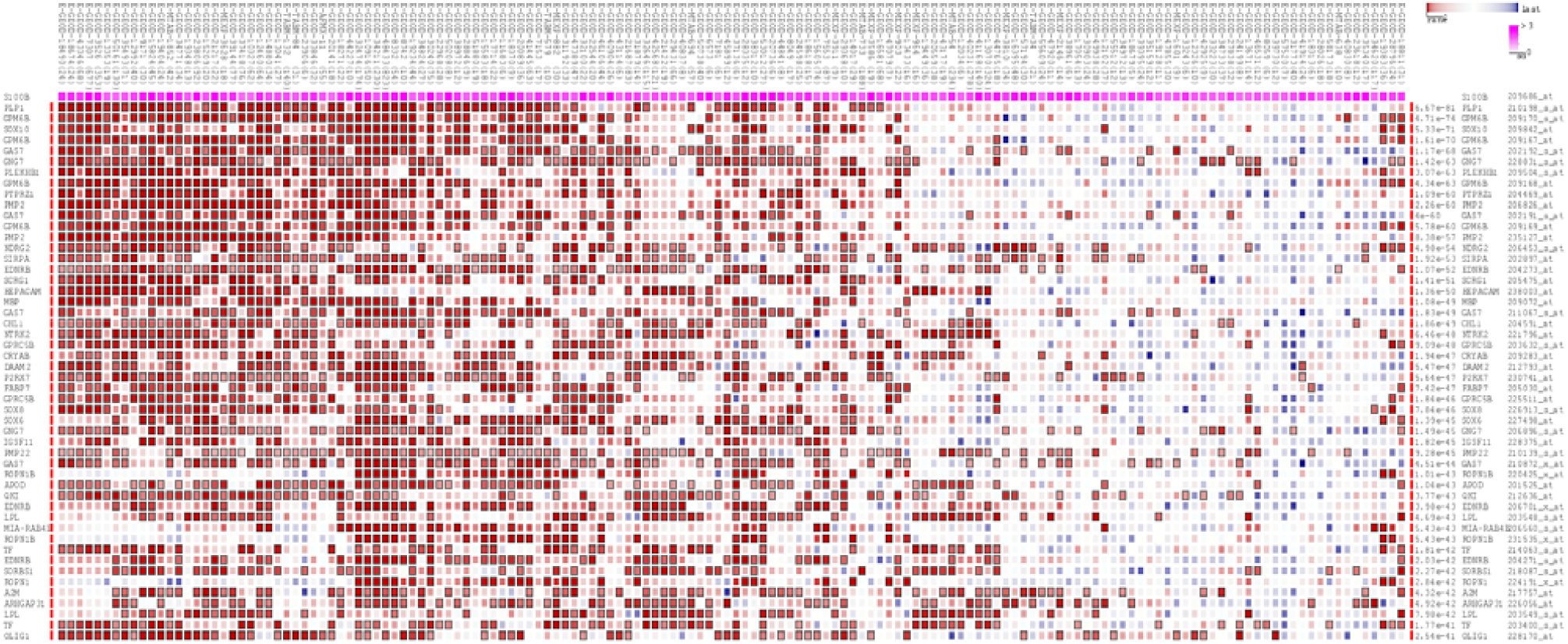
The top 150 predicted target genes of S100b by using Multi Experiment Matrix (MEM, http://biit.cs.ut.ee/mem/) website.

In most of the research results, S100B has a higher expression level in the serum or cerebrospinal fluid of epileptic patients than in ordinary people. This clear expression level trend provides the basis for establishing baseline standards. In the future, the clinical diagnosis of epilepsy can be based on the level of serum S100B to determine the clinical endpoint, and doctors can also use the above criteria to predict the onset of epilepsy in the future. The current evidence cannot prove that clinical practitioners can use the level of S100B as an evaluation strategy for the diagnosis or prognosis of epilepsy. Before this, two problems must get solved. First, we need to determine whether a quantitative association exists between S100B progression and event risk across populations and interventions. Second, the link between treatment effect changes in S100B and changes in clinical endpoints must be significant in the intervention trial. To our knowledge, no publications have filled these gaps in evidence.

In clinical practice, it is impossible for a single marker to answer such questions. According to the report, the S100B level before and after the seizure is different within a few hours (1h, 3h, 8h, 24h, 48h, 72h), so the variability of the progression of the S100B level is substantial. Another limitation is the observation conditions. S100B level inspection is currently not a routine detection item for epilepsy in hospitals. Only clinical research on S100B can undergo S100B detection. To study these problems, we have developed a questionnaire and sent emails to the teams that have made reports on epilepsy related to the S100B level. The couples willing to provide detailed clinical information will become our cooperation group. This mode of investigation provides us with potentially significant and clinically complicated samples for objective evidence.

## 2 Methods

### 2.1 Study design and study population

PROG-S100B is a multinational, multi-centre project (registration: https://www.crd.york.ac.uk/PROSPERO/, CRD-ID: CRD42023425431), aiming to integrate future research from large data centre S100B The individual data from the study were combined to answer a series of questions and to resolve the debate about whether S100B levels have clinical value in epilepsy. We tried to contact the heads of epilepsy research centres worldwide researching S100B and epilepsy-related epilepsy to cooperate more efficiently to complete this research. Investigators are collaborating to build an extensive S100B database. With the dynamic operation of bibliometrics, new reports on S100B and epilepsy-related research will gradually appear so that this project will incorporate more data sources.

### 2.2 Patient enrollment and study conduction

The first phase is mainly observational studies, focusing on resolving the debate on the correlation between S100B levels in serum or cerebrospinal fluid and epilepsy. The second phase uses observational and randomized controlled research data, focusing on solving the baseline standard of S100B level and epilepsy population with different characteristics and providing a research basis for the predictive value of S100B level on epilepsy. The third phase is mainly based on randomized controlled studies, focusing on the accuracy of S100B as a biomarker for diagnosing epilepsy. Regarding the three-stage tasks of studying epilepsy and S100B, the series of problems we need to solve are as follows:

1. Are S100B concentration levels in serum or CSF associated with epilepsy (significantly)?
2. What is the range of baseline levels of S100B concentrations in serum or cerebrospinal fluid in different types of healthy people?
3. What is the range of baseline levels of S100B concentration levels in serum or cerebrospinal fluid in epilepsy patients with different characteristics or high-risk epilepsy groups?
4. How to transform (2) and (3) into a set of feasible solutions to provide predictive value for clinical epilepsy patients?
5. Do S100B concentration levels in serum or CSF serve (significantly) as a diagnostic biomarker for epilepsy?
6. Can S100B concentration in serum or CSF be quantified as a prognostic marker for epilepsy treatment?

To solve the above six problems in the three-stage tasks, we mainly searched the original research on epilepsy-related S100B in PubMed (**Supplementary Table 1**). We searched for the contact information of the person in charge of the epilepsy research centre of epilepsy related S100B worldwide through the search results. As well as the company that produces the S100B detection kit, if necessary, it is essential to search the reference list of the above search results manually.

For the multi-centre, multi-country large-sample meta-analysis, the best strategy for collecting data is to find the heads of epilepsy research centres that study S100B and then convince these heads to cooperate with us, which can not only avoid extracting data from published articles. It can also become a research model for meta-analysis and systematic review. In addition, the epilepsy research team that studies S100B-related epilepsy has advantages over recruiting epilepsy research teams. These advantages are mainly reflected in the fact that it is easier to recruit epilepsy patients and has more weight in academic research. Once the person in charge is willing to cooperate with us, we will nominate the person in the direction to enter the program-S100B project after signing documents such as informed consent.

The study had been run continuously for one year before the publication of this study protocol. We have developed inclusion criteria (**Table 1**), but slight changes may be made when the project is run. After the PubMed search, we made the search results of epilepsy and biomarkers into a visualized chart(4), which is convenient for understanding the latest research trends in the world, and the 37 research results that meet our project have been compiled into the list of research that planned to be recruited (**Supplementary Data**).

**Table 1.** Inclusion criteria for stages 1 to 3 of the PROG-S100B project.

| Stage 1 | Stage 2 | Stage 3 |
| --- | --- | --- |
| Prospective longitudinal study design |  |  |
| Observational study |  | Randomized controlled trial |
| ○The experimental group consisted of patients diagnosed with epilepsy<br>○The control group consisted of healthy people | Investigation of one of the following at-risk populations:<br>-Malaria<br>-Neural cysticercosis<br>-Brain injury<br>-Brain deformity |  |
|  | -Meningitis<br>-Encephalitis<br>-Brain tumor<br>-Neurogenetic disease<br>-Other defined at-risk populations if a sufficient number of studies/trials is found |  |
|  | Alternatively, population-based samples with a relevant subcohort (at least 10 events) in at least one of the above-named at-risk categories |  |
| Well-defined and disclosed inclusion criteria and recruitment strategy |  |  |
| At least two EEG and S100B serum tests were performed to determine the type of epilepsy and S100B level |  |  |
| A minimum of 10 events per endpoint |  | No minimum rate of endpoint events |

### 2.3 Analysis strategy for stage 1

We have found that etiologies of epilepsy include but are not limited to stroke, tumour, and genetics. Subjects suffering from epilepsy must meet the diagnostic criteria of WHO or the International Society Against Epilepsy or be excluded from the first stage analysis. The clinical endpoint event was seizure or non-seizure, and the starting point event was a diagnosis of epilepsy, evaluation of treatment effect and prediction of epilepsy recurrence. Included studies used different definitions of clinical thresholds. We will try to select variables with clinical starting points as consistently as possible to reduce heterogeneity.

Our research topic is whether there is a correlation between epilepsy and the concentration of S100B. Seizures are easier to judge because of convulsions, and the most critical observation index is the S100B level (serum or cerebrospinal fluid), so the S100B level will be used as the primary target for data extraction. Due to the small number of variables, explaining the differences in S100B levels in different populations is challenging. Age and region may be used as correction variables to avoid S100B measurement errors, and we can use linear regression to estimate the progress of individual S100B levels. We then reanalyzed the comparisons based on estimates of the mean difference between the epileptic and non-epileptic groups, correcting for population bias.

### 2.4 Analysis strategy for stage 2

The second phase of the PROG-S100B project continues the observational study of the first phase and adds high-risk groups for epilepsy. If the survey recruits a large enough sample, the baseline criteria for S100B in different high-risk groups will confirm. Inclusion criteria may get lowered if fewer people are recruited (**Table 1**).

### 2.5 Analysis strategy for stage 3

Clinical trials measuring S100B have more EEG and S100B examinations are performed more frequently. The regression model was used to test the confounding factors, and the correlation of S100B in different risk groups was analyzed. The extensive sample data is based on the previously determined S100B concentration baseline interval to determine the breakpoint value. Taking the clinician’s diagnosis as the gold standard and the breakpoint value of S100B concentration as the pseudo-diagnosis, the true positive, false positive, true negative and false negative of epilepsy were deduced. The sample sizes of the four characteristics were stratified according to the studies, and joint tabulations were made to calculate the sensitivity and 95% confidence interval of S100B for diagnosing epilepsy.

For the treatment experiments, we used the model of van Houwelingen (5,6). For diagnostic investigations, we use a model that balances accuracy and two-dimensionality (7,8). Regardless of treatment experiments or diagnostic tests, it is easier to obtain routine data processing results such as mean logit sensitivity, mean logit specificity, standard error, and 95% confidence interval. It is also necessary to calculate variability and covariance to produce visual effects. After hard work, the total receiver operating curve (ROC) of the S100B stratification study is presented in an elliptical contour diagram.

Heterogeneity across study centres was assessed using I^2^. We used funnel plots and asymmetric regression tests to rule out publication bias. To assess methodological bias, we used only one person for the assessment (Yan Bo).

### 2.6 Statistical calculations

Obtaining quantitative baseline data and confirming associations between S100B and subgroups in large high-risk epilepsy populations is complicated. The following planned data analysis strategies are necessary, but unplanned ones may appear in the actual process. Due to the study design in the enacted recruitment strategy as observational and randomized controlled studies, the data we obtained may be two-arm or single-arm.

We will use Stata’s “meta” command package to perform random effects meta-analysis using inverse variance weighting. Categorical data were used to calculate risk ratios (RRs) using dichotomous or dummy variable methods, and the rest data was transformed into continuous data to calculate the mean and standard deviation.

We then employed a Cox regression model, utilizing the method of moments approach of DerSimonian and Laird (9), to study the influence of epileptic disorder characteristics on S100B levels. To eliminate the potential effect of time on S100B levels, period variables such as years of epilepsy diagnosis, seizure frequency, and the interval between seizures and detection of S100B were used as covariates. Measuring principles and kit companies for S100B levels were considered potential sources of heterogeneity, and these variables would be appropriately added to the model. In recruiting patients in each research centre, we subjectively believe that age and gender are manipulated and may not be the primary source of heterogeneity at the S100B population level. However, considering the characteristics of high-risk groups of epilepsy, we are worried that age and gender factors are the epidemiological factors of epilepsy, so we have to add them to the model. There may be more potential confounding factors that have not been considered, and we can only add them if we have accurate data from each centre.

## 3 Discussion

### 3.1 Annual trend analysis of research trends on biomarkers of epilepsy

1767 works of literature were included in this survey of epilepsy-related biomarker research trends (**Supplementary Table 1**). From **Supplementary Figure 1**, we can see that the overall number of published papers is on the rise, and relevant research articles have risen sharply since 2017, reaching a peak (177) in 2020. In the past few years, from 2017 to 2022, the number of published articles has remained relatively high.

### 3.2 Spatial-temporal distribution and research strength of research trends on biomarkers of epilepsy

After statistical analysis, the top ten authors and their affiliated research institutions are in **Supplementary Table 2**. Among them, Dr Orrin Devinsky, whose institution is New York University, has the highest number of publications in the literature, with 24 journals.

According to the analysis results of Citespace (**Figure 2**), the author graphs of literature are scattered, and there are few connections. The number of nodes for authors and research institutions is 1046, the number of connections is 1665, and the density is only 0.003. In the author analysis, the centrality of each node is 0, indicating that the author is not closely related. They have mainly formed a team led by Orrin Devinsky.

**Figure 2.**
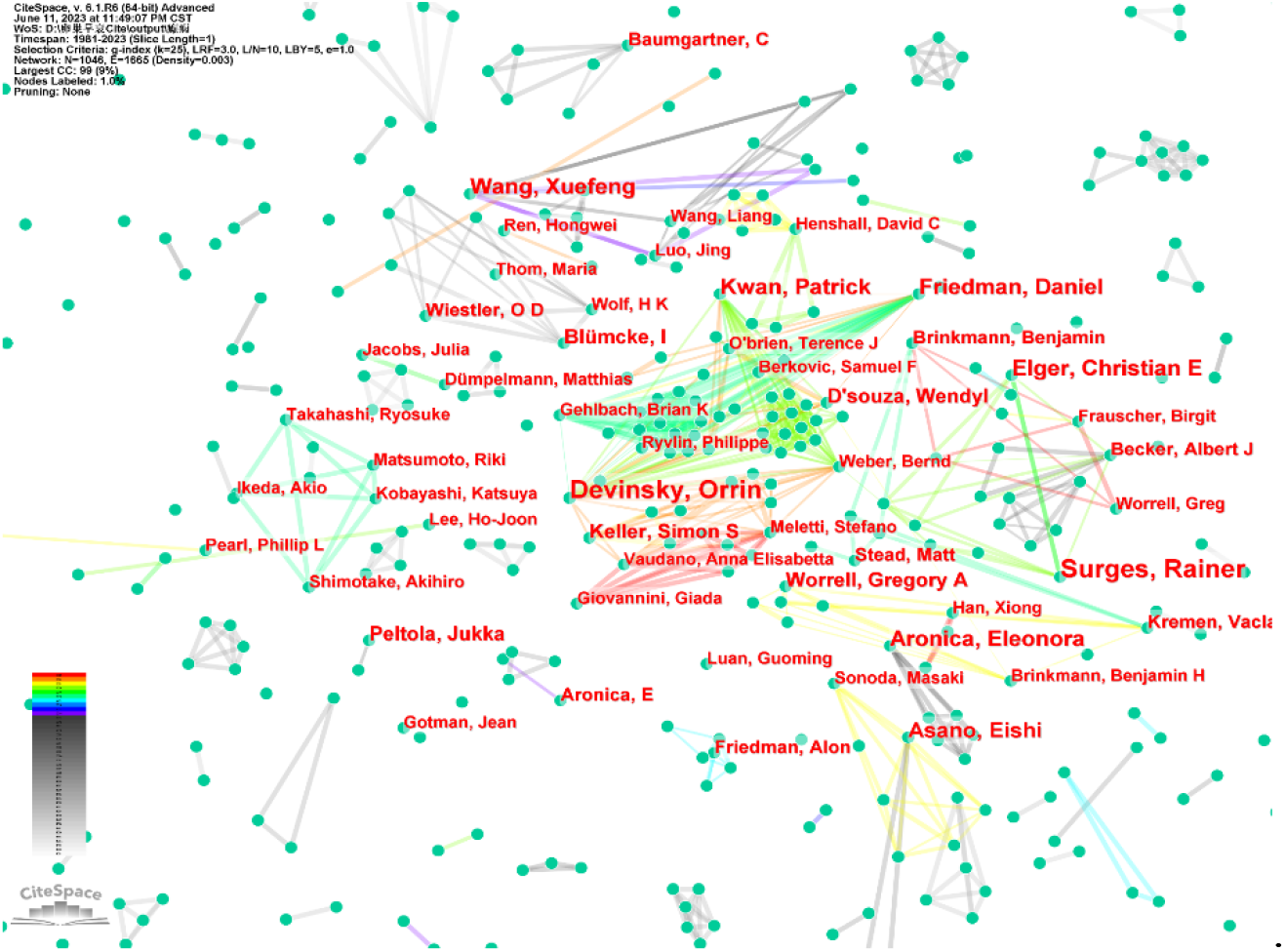
Literature author publishing cooperation network.

### 3.3 Hotspots and frontiers of research on biomarkers of epilepsy

In this study, the keywords in the pieces of literature were statistically analyzed and sorted according to frequency. The final top ten keywords are shown in **Supplementary Table 3**. The top ten keywords with frequency include “temporal lobe epilepsy”, “epilepsy surgery”, “high-frequency oscillation”, “antiepileptic drug”, “drug-resistant epilepsy”, “oxidative stress”, “hippocampal sclerosis”, “high-frequency oscillation”, “magnetic resonance imaging”, “machine learning”.

The research trend map of biomarkers of epilepsy diseases drawn through keyword co-occurrence analysis (**Figure 3**) shows that the larger the size of the node, the greater the weight of the keyword and the more times it appears. The top 10 keywords of document centrality are “temporal lobe epilepsy”, “antiepileptic drug”, “drug-resistant epilepsy”, “epilepsy surgery”, “magnetic resonance imaging”, “hippocampal sclerosis”, “oxidative stress”, “machine learning”, “autoimmune encephalitis”, “heart rate variability”. The figure shows that the connections between keyword nodes are crisscrossed, indicating that these are closely related.

**Figure 3.**
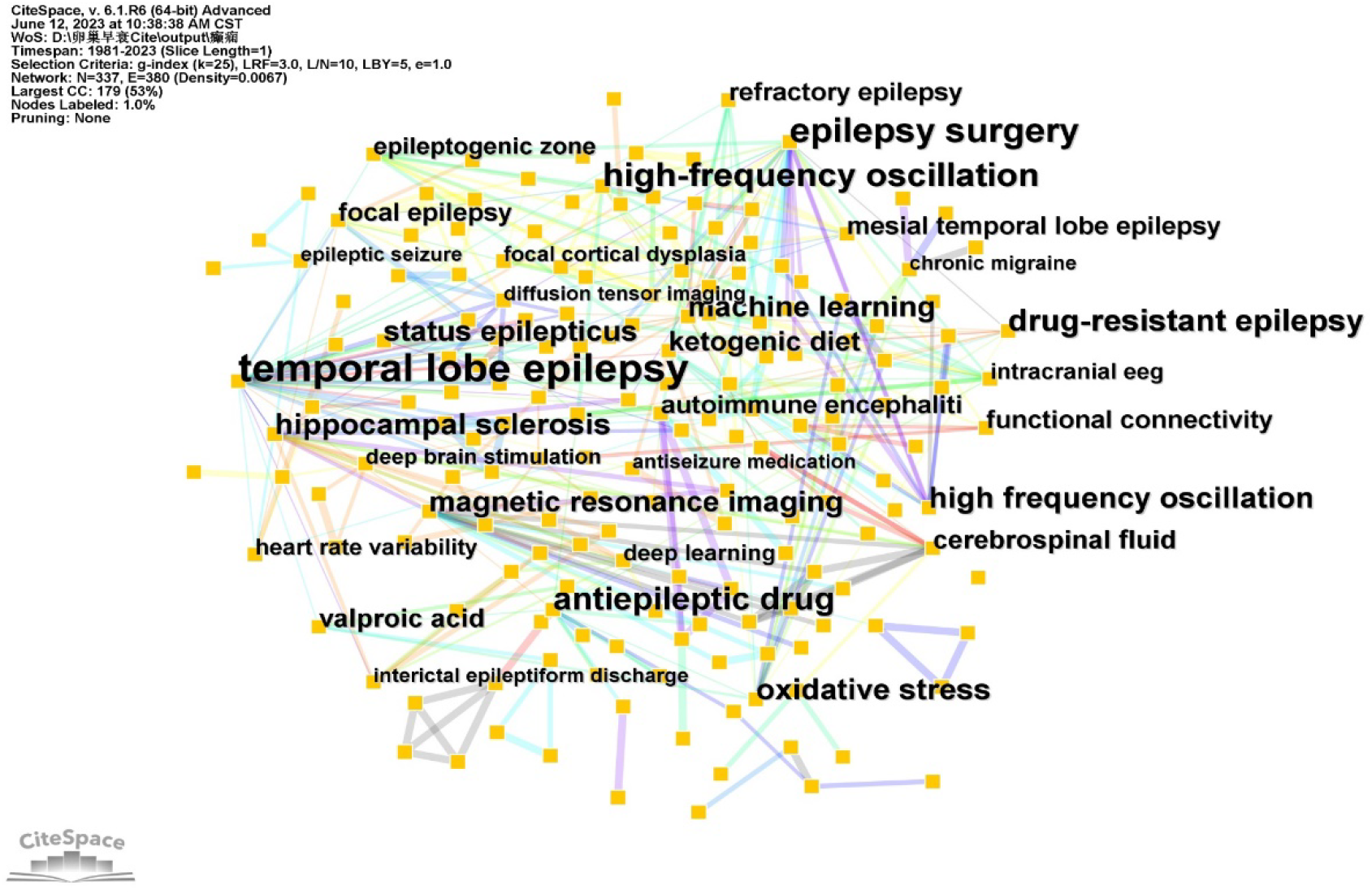
Keyword co-occurrence map of research trends in biomarkers of epilepsy.

We can find research hotspots in the current field through keyword clustering network analysis. CiteSpace software can carry out K-means clustering analysis on literature keywords to obtain clustering analysis, respectively (as shown in **Figure 4**). Among them, the number size of the cluster is inversely proportional to the range of the bunch. It can be seen from the figure that the research topics related to treatment since 1981, for #0 epileptogenic zone, #1 blood serum, #2 hippocampal sclerosis, #3 cerebrospinal fluid marker, #4 antiepileptic drug, #5 oxidative stress, #6 methylation status. From the clustering, we can notice that the current common attributes are mainly concentrated in cerebrospinal fluid markers.

**Figure 4.**
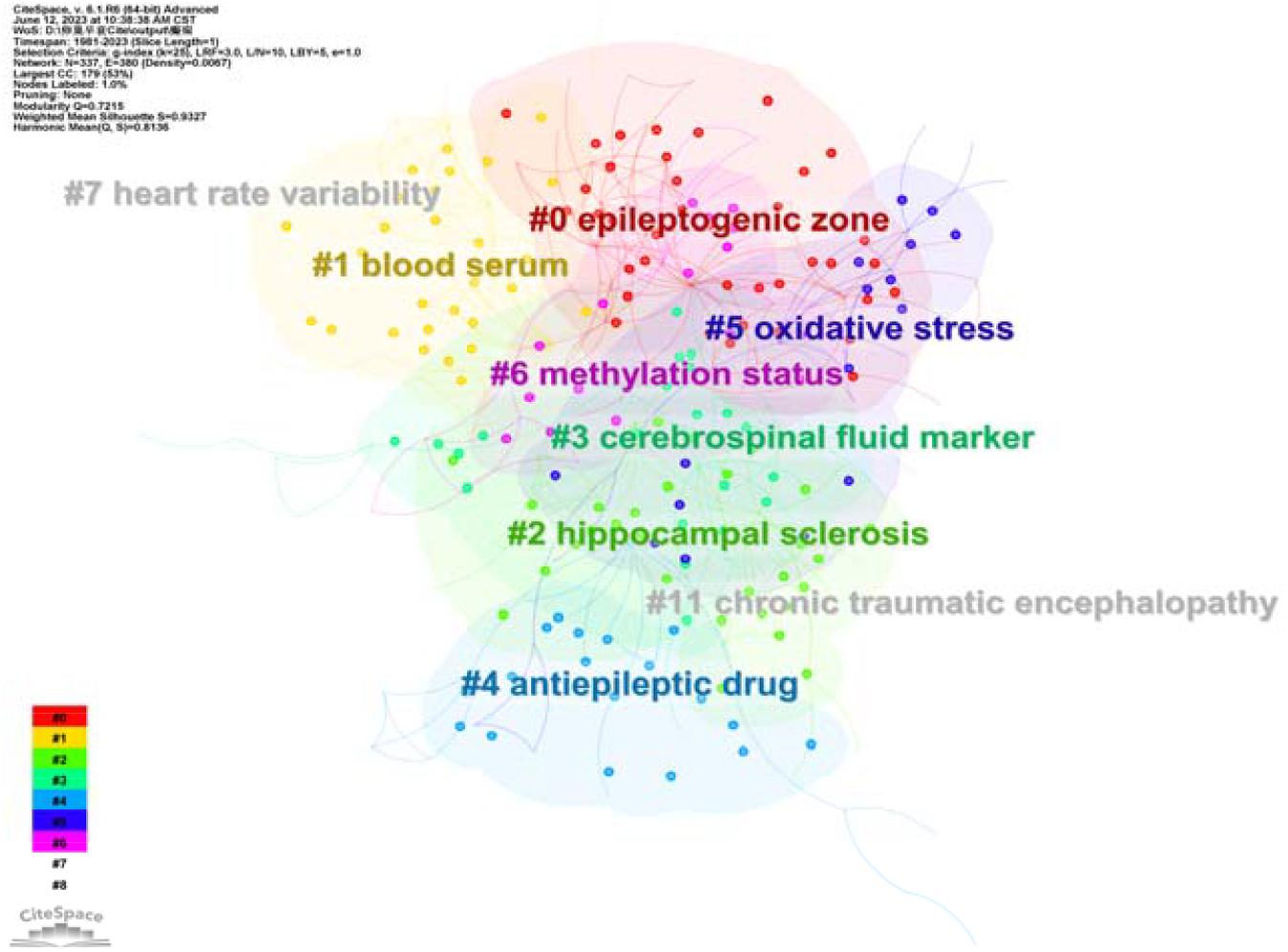
Research Trends of Epilepsy Biomarkers Keyword Cluster Analysis.

## 4 Summary

The program-S100B project addresses the relationship between changes in S100B over time and the risk of clinical events in epilepsy. Given the complex nature of the data available, this project required extensive statistical analysis. This project required the collaboration of many colleagues in the field of epilepsy research and is a normative research initiative in epilepsy.

## Data Availability

All data produced in the present study are available upon reasonable request to the authors

https://www.crd.york.ac.uk/PROSPERO/view/CRD42023425431

## 6 Acknowledgments

We are grateful Haodong Yu from Guangming District People’s Court of Shenzhen in China for writing the legal and ethical documents such as the informed consent form, Ren Sha for managing the revenue and expenditure of this study to provide sound financial decisions to maintain the normal operation of the study, Wenjie Jiang from Northwest Minzu University for bibliometrics to keep abreast of the latest research progress of epilepsy biomarkers in real time, Prof. Dr. Liyang Ma and Prof. Dr. Fei Zhao from Northwest Minzu University for the data processing support, the Guangming District People’s Court of Shenzhen in China for its legal and regulatory support.

## 10 Supplementary Data

### 10.1 Recruitment list of the PROGS100B Study Group

Dr Johan Zelano

Department of Clinical Neuroscience, Sahlgrenska Academy, University of Gothenburg, Sweden.

Dr Parisa Gazerani

Department of Health Science and Technology, Faculty of Medicine, Aalborg University, Aalborg, Denmark

Dr Geir Bjørklund

Council for Nutritional and Environmental Medicine, Toften 24,8610 MoiRana, Norway

Dr Elzbieta Bronisz

Second Department of Neurology, Institute of Psychiatry and Neurology, 02-957 Warsaw, Poland

Dr Iwona Kurkowska-Jastrzębska

Department of Neurology, Institute of Psychiatry and Neurology, Warsaw, Poland

Dr Jihong Tang

Department of Neurology, Children’s Hospital of Soochow University, No.92, Zhongnanjie Road, Suzhou 215025, Jiangsu, China

Dr Yao-Chung Chuang

Department of Neurology, Kaohsiung Chang Gung Memorial Hospital and Chang Gung University College of Medicine, Kaohsiung, Taiwan

Dr Fang Wen

Department of Neurology, Pukou Branch Hospital of Jiangsu Province Hospital (Nanjing Pukou Central Hospital), Nanjing, Jiangsu Province, China

Dr Christian Geyer

Department of Pediatric Surgery, University of Leipzig, Liebigstrasse 20a, 04103 Leipzig, Germany

Dr Xiaoming Liu

Department of Neurology, Xuzhou Children’s Hospital, Xuzhou Medical University, Xuzhou, Jiangsu 221000, P.R. China

Dr Yuping Wang

Epilepsy Centre, Department of Neurology, Xuanwu Hospital, Capital Medical University, 45 Chang Chun Street, Beijing 100053, China.

Dr Johanna Palmio

Department of Neurology, University of Tampere, FIN-33014 Tampere, Finland

Dr Mustafa Calik

Harran University School of Medicine, Department of Pediatric Neurology, Sanliurfa 63100, Turkey

Dr W Sue T Griffin

Department of Neurobiology and Developmental Sciences, University of Arkansas College of Medicine, Little Rock, AR 72205, USA

Dr Luis V. C.

Portela, Departamento de Bioqumica, ICBS, UFRGS. Rua Ramiro Barcelos 2600, anexo, Porto Alegre, RS, Brazil. CEP: 90035-003

Dr Yun Xu,

Department of Neurology, Drum Tower Hospital of Nanjing Medical University, 321 Zhongshan Road, Nanjing Jiangsu 210008, China.

Dr Mustafa Calik

School of Medicine, TR-63100 Sanliurfa, Turkey

Dr Stefano Meletti

Department of Biomedical, Metabolic, and Neural Sciences, Center for Neurosciences and Neurotechnology, University of Modena and Reggio Emilia, Modena, Italy.

Dr Vincent Navarro

Sorbonne Université, Institut du Cerveau – Paris Brain Institute – ICM, Inserm, CNRS, APHP, Hôpital de la Pitié-Salpêtrière, DMU Neurosciences, Paris France

Dr Gang Yu

The First Affiliated Hospital of Chongqing Medical University, No.1, Youyi Road, Yuanjiagang, Yuzhong District, Chongqing 400000, China

Dr Kirsi Mikkonen

Department of Pediatrics, University of Oulu, P.O. Box 5000, FIN-90014 University of Oulu, Finland

Dr Monika Mochol

Department of Neurology, Østfold Hospital Trust, Norway

Dr Mohamed Khamis

Neurology, Ain Shams University, Cairo, Egypt

Dr Elzbieta Bronisz

Second Department of Neurology, Institute of Psychiatry and Neurology, 02-957 Warsaw, Poland

Dr Luis V.C. Portela

Departamento de Bioquímica, ICBS, Universidade Federal do Rio Grande do Sul, Avenida Ramiro Barcelos, 2600-Anexo, CEP 90035-003, Porto Alegre, RS, Brazil;

Dr Suvi P. Liimatainen

Department of Neurology and Rehabilitation, Tampere University Hospital, P.O. Box 2000, 33521 Tampere, Finland

Dr Leila Simani

Skull Base Research Center, Loghman-Hakim Hospital, Shahid Beheshti University of Medical Sciences, South Kargar Ave, Kamali St, Tehran, Iran.

Dr Xueyuan Liu

Department of Neurology, (Shanghai Tenth People’s Hospital), Tenth People’s Hospital of Tongji University, No. 301 Middle Yanchang Road, Shanghai 200072, China

## 11 Supplementary Figures and Tables

### 11.1 Supplementary Tables

**Supplementary Table 2.**
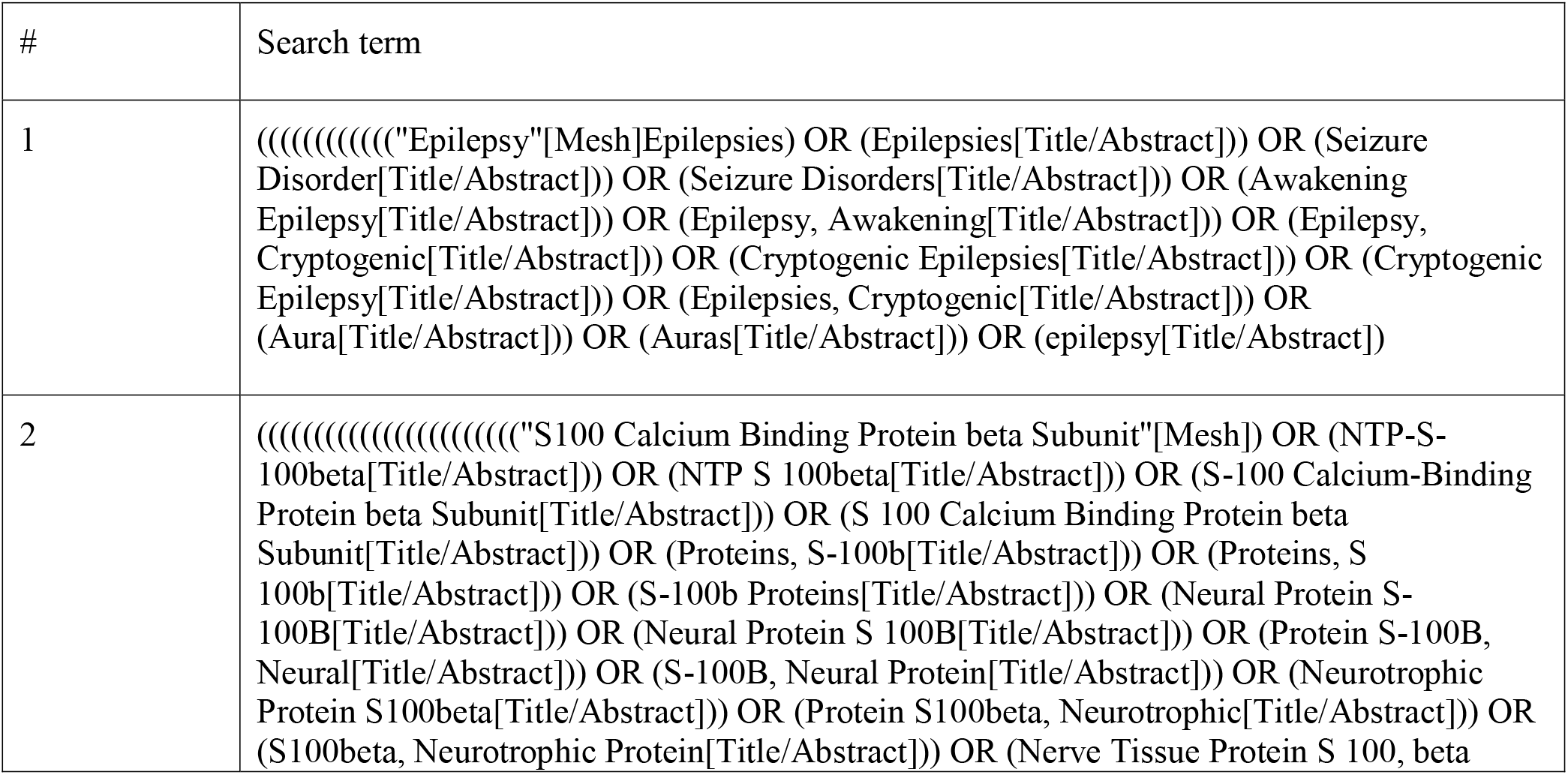
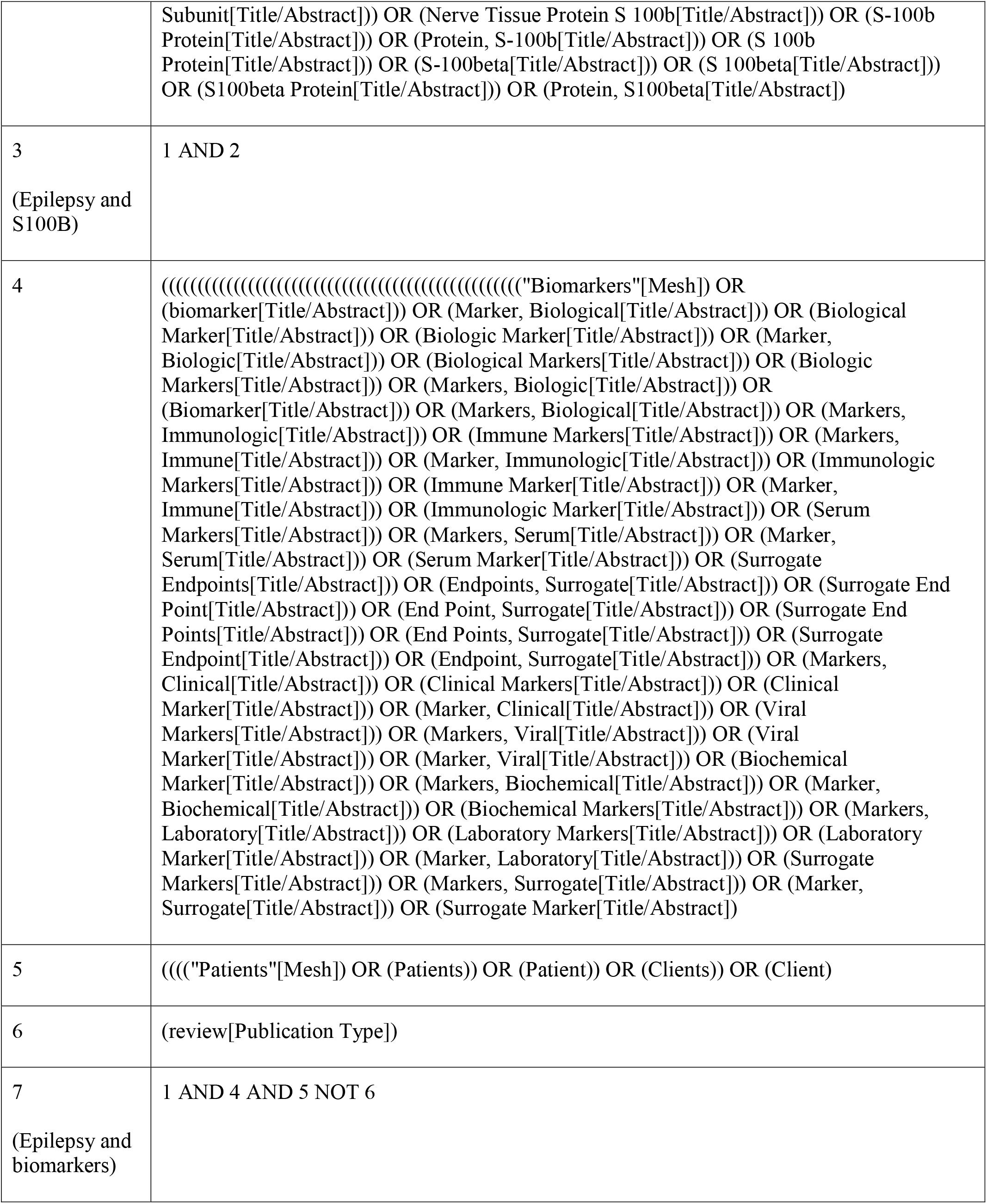
The full search strategy.

**Supplementary Table 3.** Distribution of the top ten authors of research trends on biomarkers of epilepsy.

| Rank | Author | Frequency | institution |
| --- | --- | --- | --- |
| 1 | Orrin Devinsky | 24 | New York University |
| 2 | Eleonora Aronica | 18 | Academisch Medisch Centrum Universiteit van Amsterdam \| AMC · Department of Pathology |
| 3 | Patrick Kwan | 16 | Monash University, Department of Neuroscience |
| 4 | Daniel Friedman | 16 | New York University Langone School of Medicine |
| 5 | O D Wiestler | 14 | Department of Pathology, University of Zürich, Switzerland |
| 6 | Stefano Meletti | 12 | Department of Biomedical, Metabolic and Neural Sciences, Center for Neuroscience and Neurotechnology, University of Modena and Reggio Emilia, Modena, Italy; Department of Neurosciences, OCSAE Hospital, AOU Modena, Modena, Italy. |
| 7 | Anna Elisabetta Vaudano | 12 | Department of Biomedical, Metabolic, and Neural Science, University of Modena and Reggio Emilia, Modena, Italy; N.O.C.S.A.E. Hospital, ASL Modena, Italy. |
| 8 | Simon S Keller | 12 | The Magnetic Resonance and Image Analysis Research Centre (MARIARC), United Kingdom. |
| 9 | Wendyl D'Souza | 12 | Medicine - St Vincent's Hospital |
| 10 | Ingmar Blumcke | 12 | Institute of Neuropathology, University Hospitals Erlangen, Erlangen, Germany. |

**Supplementary Table 4.** Research Trends Keyword Analysis of Biomarkers for Epilepsy Diseases (sorted by frequency).

| Rank | Frequency | Centrality | Keywords |
| --- | --- | --- | --- |
| 1 | 59 | 0.23 | temporal lobe epilepsy |
| 2 | 35 | 0.1 | epilepsy surgery |
| 3 | 29 | 0.03 | high-frequency oscillation |
| 4 | 26 | 0.11 | antiepileptic drug |
| 5 | 22 | 0.11 | drug-resistant epilepsy |
| 6 | 21 | 0.06 | oxidative stress |
| 7 | 20 | 0.07 | hippocampal sclerosis |
| 8 | 20 | 0.02 | high frequency oscillation |
| 9 | 18 | 0.1 | magnetic resonance imaging |
| 10 | 17 | 0.05 | machine learning |

**Supplementary Table 4.** Research Trends Keyword Analysis of Biomarkers for Epilepsy Diseases (sorted by centrality).

| Rank | Frequency | Centrality | Keywords |
| --- | --- | --- | --- |
| 1 | 59 | 0.23 | temporal lobe epilepsy |
| 2 | 26 | 0.11 | antiepileptic drug |
| 3 | 22 | 0.11 | drug-resistant epilepsy |
| 4 | 35 | 0.1 | epilepsy surgery |
| 5 | 18 | 0.1 | magnetic resonance imaging |
| 6 | 20 | 0.07 | hippocampal sclerosis |
| 7 | 21 | 0.06 | oxidative stress |
| 8 | 17 | 0.05 | machine learning |
| 9 | 11 | 0.04 | autoimmune encephaliti |
| 10 | 9 | 0.04 | heart rate variability |

### 11.2 Supplementary Figures

**Supplementary Figure 1.**
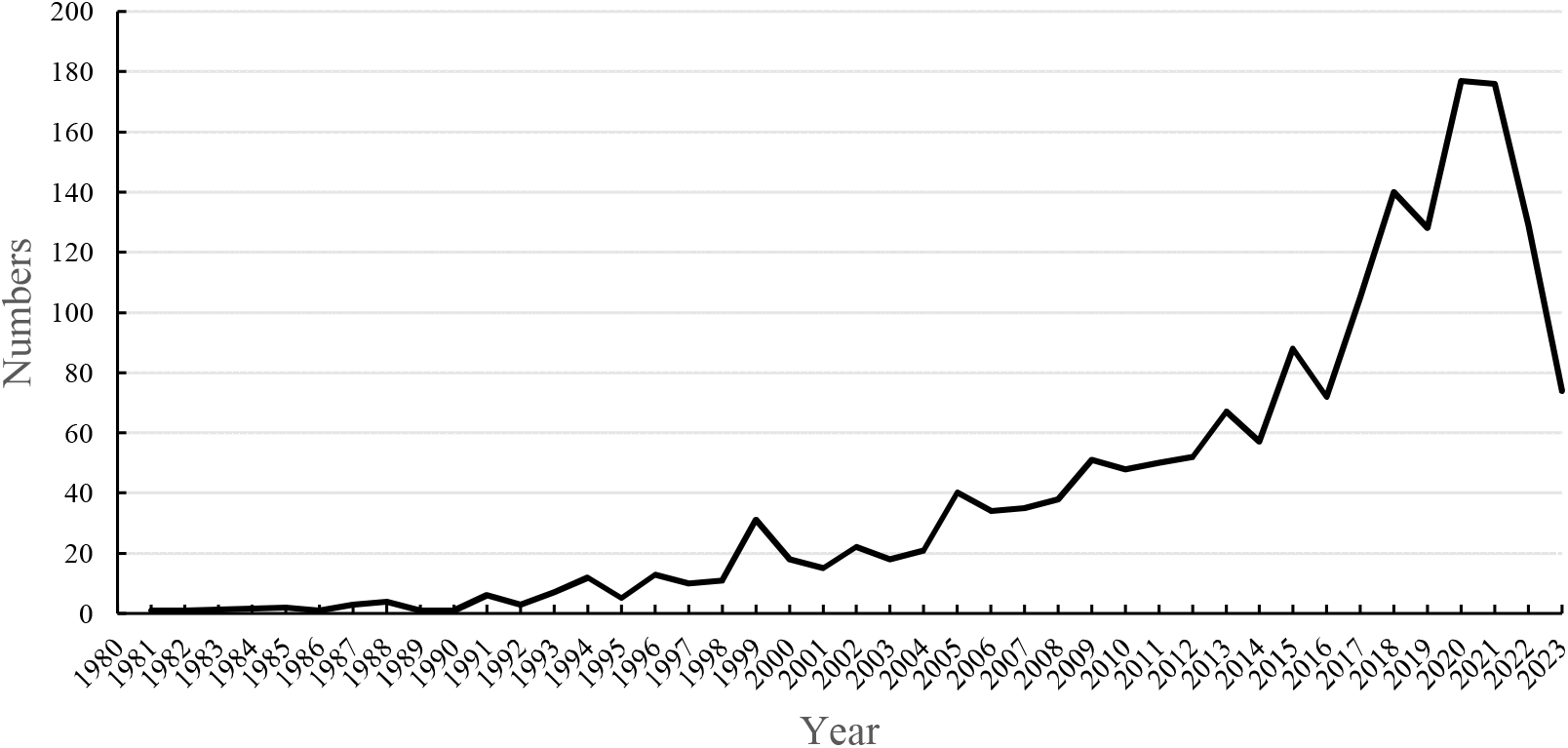
Trends in literature publishing years.

